# Long term smoking and quitting among people with severe mental illness: 3-year follow-up of the SCIMITAR+ Trial

**DOI:** 10.1101/2024.06.03.24308386

**Authors:** Simon Gilbody, the SCIMITAR trials collaborative

## Abstract

**Background:** People with severe mental illnesses (SMIs) are three times more likely to smoke than the wider population, contributing to widening health inequalities. Here we report the long term [3 year] outcomes for the SCIMITAR+ trial (ISRCTN72955454), which compared usual care to a bespoke smoking cessation package.

**Methods:** We recruited 526 heavy smokers with bipolar illness or schizophrenia who were randomly allocated to a bespoke smoking cessation intervention (n=265) or to usual care (n=261) between October 2015 and December 2016. We measured long-term quitting by carbon monoxide-verified smoking status at 3 years post randomisation, and a range of secondary outcomes.

**Results:** 261 of the original 526 were followed up. 209 participants (80.1% of followed-up; 39.7% of randomised) could be defined as a smoker or non-smoker for the 36-month primary analysis. Forty-three participants (16.5% of 261) were determined to be a non-smoker via self-reporting and CO confirmation; 21 in the intervention arm (16.3% of 129; 19.6% of those who provided both self-reported and CO measure) and 22 in the control arm (16.7% of 132; 21.6% of those who provided both self-reported and CO measure) – adjusted OR of 0.89 (95% CI 0.45, 1.77, p = 0.74).

For secondary outcomes, there was no sustained between group difference in reduction in nicotine dependence (measured using the Fagerstrom Test for Nicotine Dependence) or motivation to quit. Some short-term improvements in physical health (measured by the Short Form 12) were present at 36 months.

**Conclusions:** A bespoke intervention represents the model for care for mental health services in the UK, but long term quit rates cannot be assumed. Sustained attention to smoking and relapse is likely to be needed to ensure that short term gains are maintained. The certainty of these results is tempered by loss to follow up and low statistical power.

**Funding:** This study was funded by NIHR Health Technology Assessment Programme (Project number 11/136/52)

**Plain English Summary:** Smoking rates are very high amongst people who use mental health services. This makes a significant contribution to health inequalities and reduced life expectancy. Earlier research has shown that smoking cessation services are effective when the specific needs of people with mental ill health are taken into account. This forms the basis of guidelines issued by the National Institute for Health and Care Excellence. In this study we looked at the longer-term impact of an intervention to help people to quit smoking. At three years under half of the people who took part in the original study were able to provide information on their use of tobacco. The rates of smoking were similar between people who received the intervention three years ago, and those who received only usual care. Since we already know that the intervention is effective in helping people to quit in the short term, it would be prudent to ensure that people continue to be offered care to help them to quit if they are at risk of relapse. The longer-term impact of a brief intervention on longer term quit rates cannot be assumed, since the strength of this conclusion is limited because we were only able to follow up under half of the people who took part in the study. Further research is needed into the optimum way in which the health benefits of quitting can be sustained in the longer term.

## Introduction

People with severe mental illness (SMI), such as schizophrenia and bipolar disorder are more likely to smoke and to smoke more heavily than the wider population.^1,2^ The point prevalence of smoking amongst those with severe mental illness has been estimated to be between 58% and 90%.^3^ People with severe mental illness begin smoking at an earlier age and at a higher rate compared to smokers without severe mental illness.^4,5^ Smokers with severe mental illness smoke each cigarette more intensely, extracting more nicotine per cigarette,^6^ are more nicotine-dependent, more likely to develop smoking-related illness, and less likely to receive help in quitting, compared with the wider population of smokers.^7^

Smoking is part of the ‘culture’ of mental health services; both amongst staff and patients.^8^ This may be part of why mental health services have not historically made greater efforts to develop and deliver better smoking cessation treatments for this group. Many believe smoking relieves depression and anxiety,^9^ when the opposite is true.^10^ Smoking contributes to the general poor physical health of those with severe mental illness. Cohort studies have shown that people with severe mental illnesses such as schizophrenia die on average 20-25 years earlier than those without severe mental illnesses, and that smoking is the most important modifiable risk factor for this health inequality.^11^

Evidence derived from small scale trials suggests that people with severe mental illness are able to give up smoking and that behavioural and pharmacological intervention to aid quitting might be as effective for people with severe mental illness as for the wider population.^12^ However, people with severe mental illness do not generally access generic smoking cessation services when they have been offered in the NHS.^13^ In the general population, the rate of smoking is falling whereas there has been little shift amongst people with severe mental illness.^14^

To address this widening health inequality we designed a smoking cessation intervention specifically for people with severe mental illness, based on evidence-supported behaviour change techniques and pharmacotherapy.^15,16^ We have previously reported the short and medium term results of our SCIMITAR pilot trial,^17^ and a subsequent large scale multisite pragmatic trial know as SCIMITAR+.^18^

Analysis of these trials shows that bespoke smoking cessation,^19^ designed to meet the needs of people with severe mental illness is effective in ensuring quitting at 12 months [odds ratio of quitting OR = 1.67, 95% CI 1.02–2.73, P = 0.04)] and that it is cost effective.^20^

There are few long term follow up studies of smoking cessation studies in the field of mental health^21,22^and the SCIMITAR+ trial included a long term follow up study to capture the enduring impact of a single evidence based smoking cessation intervention. Here we report long term [36 month] follow up of our primary outcome of interest, verified quitting. To our knowledge this is the first such study to report longer term outcomes.

## Methods

### Study design and participants

In the ‘Smoking Cessation Intervention for Severe Mental Illness’ (SCIMITAR+) trial we sought to establish the clinical effectiveness of a bespoke smoking-cessation intervention compared with usual care for people with severe mental illness. SCIMITAR+ was a two-arm parallel group individually randomised controlled trial (RCT), which used a pragmatic design.^23^ The protocol to the SCIMITAR+ trial has previously been published,^24^ and the details of the trial were prospectively registered (ISRCTN72955454). Ethical approval was granted (NRES Committee Yorkshire & The Humber - Leeds East REC, 19/03/2015, ref: 15/YH/0051). The full design details have been reported previously.^18^

Inclusion criteria: aged 18 and above, have a severe mental illness, be living in the community, smoke at least five cigarettes per day and express an interest in cutting down or quitting smoking. There is no agreed definition of severe mental illness so we adopted a pragmatic definition used in UK primary care,^25^ i.e. a documented diagnosis of schizophrenia or delusional/psychotic illness (corresponding with categories F20·X & F22·X from the 10th revision of the International Classification of Diseases - ICD 10) or bipolar disorder (ICD 10 F31·X). This severe mental illness- inclusive diagnosis needed to have been made by specialist mental health services and documented in either the primary-care records or psychiatric notes prior to recruitment. We excluded people who were pregnant or breastfeeding, had significant co-morbid drug or alcohol problems (as ascertained by the GP or mental health worker), were non-English speakers, or who lacked capacity. People in inpatient settings at the time of recruitment were excluded. Participants were made aware of the trial by members of the clinical team (either face to face or by personalised letter of invitation).

### Randomisation and masking

Simple randomisation was used following a computer-generated random number sequence. To ensure concealment of allocation, a secure telephone randomisation service run by the York Trials Unit was used. Researchers phoned the service once the participant had consented and completed the baseline assessment. Once given the details of the participants’ allocation, the researcher immediately informed them of their allocation. Due to the nature of the intervention, it was not possible to blind participants, mental health staff and primary care physicians to the treatment allocation. Researchers conducting follow up assessments were blinded to treatment allocation.

### Procedures

All participants in the trial received usual care and had access to the full range of smoking cessation treatments that were offered by their local NHS services. Under usual care, people with severe mental illness were able to access smoking cessation services provided by their primary care physician or in a locally-provided service not specifically designed for people with severe mental illness, at no direct cost. They were also able to access a free telephone helpline (‘Smokefree National Helpline’’) offering smoking cessation advice. All participants remained under the care of their primary care physician and continued to receive their usual service from the mental health team throughout the trial.

#### Intervention

Participants allocated to the bespoke smoking cessation group were offered a structured smoking cessation intervention delivered by a trained mental health smoking cessation practitioner (MH-SCP). The MH-SCPs were generally experienced mental health nurses who worked in conjunction with the participant and the participant’s primary care physician or mental health specialist to provide an individually-tailored smoking cessation service. The MH-SCP intervention was delivered according to the Manual of Smoking Cessation (developed by the National Centre for Smoking Cessation Training – NCSCT - in the UK)^26^ with a number of adaptations to cater for people with severe mental illness. These adaptations included: making several assessments prior to setting a “quit date”, offering nicotine replacement prior to setting a quit date (‘cut down to quit’),^15,27^ recognising the purpose of smoking in the context of a person’s mental illness, providing home visits, providing additional face-to-face support following an unsuccessful quit attempt or relapse, and informing the primary care physician and psychiatrist of a successful quit attempt, such that they can review anti-psychotic medication doses if metabolism changes. The MH-SCPs were drawn from local NHS staff and attended a two-day training event based on the National Centre for Smoking Cessation Training’s practitioner training with some additional training on the specific adaptations for people with severe mental illness. The MH-SCP advised on the range of pharmacological aids smoking cessation (such as nicotine replacement, and varenicline) and liaised with the participant’s primary care physician to ensure that these were offered in line with patient choice. The full range of nicotine replacement and smoking cessation products from the British National Formulary were made available to participants.^28^ However, the final prescription of treatments was left within the discretion of the primary care physician. Participants were offered up to 12 individual face to face sessions in their home or NHS premises lasting approximately 30 minutes. The intervention had been developed and tested in the context of a pilot RCT and the full details have been published elsewhere.^16,17^ Participants were contacted and the treatment programme initiated within seven days of randomisation.

#### Control

SCIMITAR+ was a pragmatic trial,^23^ and the comparator was the care that patients with severe mental illness would access under usual circumstances (‘usual care’). In the UK all patients (including people with severe mental illness) have access to statutory smoking cessation services at no direct personal cost, and this would include access to a smoking cessation counsellor delivering evidence-supported treatments, including behavioural support and access to pharmacotherapy.Participants allocated to the control group were advised to quit, to see their GP and to contact local NHS stop smoking services. Thereafter, no additional treatment was offered in the context of the SCIMITAR+ trial.

### Outcome measures

The **primary follow up for the primary outcome of interest** was smoking cessation at 12 months post-randomisation. A successful quitter was defined as someone with a CO measurement below 10 parts per million ^29^ and who reported that they had not smoked (‘not even a puff’) when asked about smoking in the past week (i.e. 7-day point prevalence abstinence at 12 months with CO<10ppm). In this analysis we report the extended follow up of confirmed quitting [measured in the same way] at 36 months, in line with our planned follow up and analysis.

**Secondary outcomes** at 36 months were: biologically-verified smoking cessation at six months, number of cigarettes smoked per day, Fagerstrom Test for Nicotine Dependence (FTND),^30^ Motivation to Quit (MTQ) questionnaire,^31^ Patient Health Questionnaire 8 (PHQ-8) [a shortened version of the PHQ9, omitting a question on self-harm], General Anxiety Disorder 7 (GAD-7),^32^ 12-Item Short Form Health Survey (SF-12),^33^ and body mass index (BMI).

### Sample size calculation

Results from the SCIMITAR pilot trial^17^ and earlier systematic reviews^12^ were used to inform the sample size calculation. The SCIMITAR+ (full RCT) was powered at 80% to detect a relative increase in quitting of 1.7 assuming a control quit rate of 20%, equal randomisation, and a two-sided alpha of 0.05. Allowing for 20% loss to follow up at 12 months [the primary outcome point] required a total of 393 participants to be recruited and randomised. We therefore proposed to conservatively recruit 400 participants overall, which we exceeded. The sample size of 36-month follow-up was limited to that of the initial trial, 526 participants were randomised into the trial (265 BSC, 261 UC). 450 participants returned the 12-month questionnaire (227 BSC, 223 USC). All participants who remained eligible were recontacted for inclusion in the 36-month follow-up.

### Statistical analysis

Analyses were conducted in Stata v15. All statistical tests were two-sided at the 5% significance level. Adjustment for multiplicity was not made, as a clear primary outcome is defined, and all other outcomes serve as secondary investigations.

#### Smoking cessation and smoking status

the proportions of CO-verified quitters at 36 months are presented with an unadjusted absolute risk difference and 95% confidence interval (CI) and were analysed on an intention to treat basis via separate mixed-effect logistic regression models (for each time point) adjusted for baseline smoking severity (self-reported number of cigarettes smoked per day), with site as a random effect. The risk difference, adjusted odds ratio (OR), corresponding two-sided 95% CI and p-value for the treatment effect are presented.

As sensitivity analyses we also conducted multiple imputation to account for missing data^34^, imputed self-reported quitting where biochemically-verified quitting was not available, and assumed that people with missing data were still smoking [a ‘worst case’ approach]. For comparison we also present the 12-month data, analysed in the same way.

#### Secondary outcomes

We compared number of cigarettes smoked per day (as reported as part of the FTND) at 12 and 36 months between the two groups using a mixed-effect negative binomial regression model using the log link function, adjusted for baseline smoking severity, treatment group, time and a treatment group-by-time interaction term, and site as a random effect. Incidence rate ratios (IRRs) and their associated 95% CIs and p-values are provided. Scores for FTND, MTQ, PHQ-9, GAD-7, SF-12 physical component, SF-12 mental component scores and BMI were compared between treatment groups using a covariance pattern linear mixed model. The outcome modelled was total score at six, 12 and 36 months. Each model included as fixed effects: baseline score, baseline smoking severity, treatment group, time and a treatment group-by-time interaction term, and site as a random effect. Predicted means for each group and the adjusted mean difference (with 95% CI and p-value) between treatment groups at 12 and 36 months are reported.

The PHQ-8 was used in this follow-up as opposed to the PHQ-9, which was used at 12 months, as the follow-up was done via post and this allowed us to avoid potential sensitivity topics with participants. As such, to compare the results with those from 12 months, the earlier results were recalculated omitting the question on suicidal thoughts, to give the PHQ-8. The mean scores for 12 and 36 months are presented. The adjusted mean difference at 36 months is reported and analysed as above, and in line with the equivalence of these two approaches.^35^

## Results

A total of 526 participants (265 intervention and 261 usual care) were recruited and randomised between October 2015 and December 2016 (Figure 1). The most common severe mental disorders were schizophrenia or other psychotic illness (n=343, 65.2%), bipolar disorder (n=115, 21.9%), and schizoaffective disorder (n=66, 12.5%). The majority of participants were male (58.7%), the median age was 47.2 years and participants were mostly overweight (median BMI 29.3), with long smoking histories (mean duration of smoking 29.9 years) and heavy levels of nicotine dependence (mean 24 cigarettes per day). Most participants (83.5%) felt that smoking had negatively affected their health, and 70.9% reported that they had been advised to stop smoking by their GP (see supplementary Table 1 for baseline data and demographic characteristics).

Smoking cessation at 12 and 36 months was defined as a CO measure of < 10 ppm (indicating no smoking in the last 12 hours) and self-reported cessation (indicating no smoking in the last week), with 442/526 (84.0%) providing sustained quit data at 12 months. At 36 months 261/525 (49.6% from original trial) responded to the extended follow-up.

The baseline characteristics of the participants who responded can be found in Supplementary Table 1, detailed by arm, and alongside those of the whole population who were randomised into the trial by arm. Of those followed up, there was an almost equal split of males and females (n=135 and 126 respectively), and the average age was 47.0 years (SD 11.7). From supplementary Table 1 it can be observed that overall the participants who responded at 36 months are mostly similar to the whole sample who were initially randomised into the trial, expect there appears to be less participants who stated they used recreational drugs at baseline than in the initial study.

For the extended measure of the primary outcome [Smoking cessation at 36 months, defined as a CO measure of < 10 ppm and self-reported (‘not even a puff’ in the last week)]. At three years, 210 provided a carbon monoxide reading (80.5% of followed-up; 39.9% of randomised and 260 participants (99.6% of followed up; 49.4% of randomised) provided self-reported smoking status. 209 participants (80.1% of followed-up; 39.7% of randomised) provided both and as such can be defined as a smoker or non-smoker for the primary analysis.

54 of the 261 participants (20.7%) stated that they had had ‘not even a puff’ when asked about smoking at 36 months, 26 (20.2% of 129) in the intervention arm, and 28 (21.2% of 132) in the control arm. Additionally, 93 had a CO measurement of less than 10ppm, which is used as to confirm smoking status – 53 (41.1% of 129) in the intervention group, and 40 (30.3% of 132) in the control arm. Ten participants self-reported to have stopped smoking but did not provide a CO measurement, and one participant self-reported to have stopped smoking, but had a carbon monoxide reading above 10ppm.

Forty-three participants (16.5% of 261; 20.6% of those who provided both measures) were determined to be a non-smoker via self-reporting and CO confirmation; 21 in the intervention arm (16.3% of 129; 19.6% of those who provided both self-reported and CO measure) and 22 in the control arm (16.7% of 132; 21.6% of those who provided both self-reported and CO measure) – risk difference -0.02, 95% CI -0.13 to 0.09.

When comparing the quit rate at three years in the two arms, there was an adjusted OR of 0.89 (95% CI 0.45, 1.77, p = 0.74). When comparing this to the results seen at six months (adjusted OR 2.4, 95% CI 1.2 to 4.7, p = 0.01) and one year (adjusted OR 1.6, 95% CI 0.9 to 2.8, p = 0.12) it can be seen that any effect from the intervention seems to dissipate over time. Details comparing the one year and three-year results can be found in Table 1.

**Table 1:** Summarises of the secondary outcomes by arm, at 12 months and 36 months, with adjusted means and group difference.

|  | One year |  | Three Years |  |  |  |
| --- | --- | --- | --- | --- | --- | --- |
|  | Intervention<br>(n=265;<br>n=227) | Control<br>(n=261;<br>n=223) | Intervention<br>(n=129) | Control<br>(n=132) | Difference<br>(95% CI) | p-value |
| **Number of cigarettes** | N=176 | N=191 | N=88 | N=86 |  |  |
| Mean (SD) | 20.2 (12.3) | 18.7 (12.1) | 18.2 (10.3) | 19.7 (10.0) | 0.92 (0.78, 1.07) | 0.27 |
| **FTND** | N=169 | N=186 | N=101 | N=98 |  |  |
| Mean (SD) | 5.6 (2.0) | 5.3 (2.3) | 5.8 (2.2) | 5.7 (1.9) | -0.00 (-0.48, 0.47) | 1.00 |
| **MTQ** | N=200 | N=200 | N=115 | N=114 |  |  |
| Mean (SD) | 13.0 (3.3) | 12.3 (3.4) | 13.1 (3.5) | 12.5 (3.6) | 0.68 (-0.10, 1.46) | 0.09 |
| **PHQ-8** | N=213 | N=211 | N=127 | N=131 |  |  |
| Mean (SD) | 8.5 (6.3) | 9.2 (6.2) | 9.0 (6.3) | 9.0 (6.5) | 0.09 (-1.14, 1.31) | 0.89 |
| **GAD-7** | N=214 | N=212 | N=128 | N=131 |  |  |
| Mean (SD) | 7.0 (6.3) | 7.4 (6.0) | 6.8 (6.1) | 7.3 (6.3) | 0.41 (-1.59, 0.77) | 0.50 |
| **SF-12, physical** | N=212 | N=207 | N=119 | N=123 |  |  |
| Mean (SD) | 44.3 (10.1) | 42.4 (11.4) | 42.2 (10.9) | 40.6 (11.7) | -0.11 (-2.34, 2.11) | 0.92 |
| **SF-12, mental** | N=212 | N=207 | N=119 | N=123 |  |  |
| Mean (SD) | 39.3 (11.9) | 38.9 (11.9) | 39.0 (13.7) | 38.2 (11.6) | -0.39 (-2.01, 1.22) | 0.64 |
| **BMI** | N=208 | N=201 | N=90 | N=80 |  |  |
| Mean (SD) | 30.4 (7.2) | 29.7 (6.7) | 31.3 (7.7) | 31.0 (6.9) | -0.03 (-1.34, 1.29) | 0.97 |

As with the analysis at one year, we conducted three separate sensitivity analyses. When imputing self-reported smoking status where CO measures were missing, analysed and adjusted in the same way as the primary, the results were similar: OR 0.88, 95% CI 0.47, 1.65, p = 0.70. When assuming that anyone with a missing smoker status (but had returned the questionnaire) was a smoker, the following results were obtained: OR 0.97, 95% CI 0.49 to 1.91, p = 0.93. Multiple chained imputation of missing data was not undertaken, due to lack of missing data.

### Secondary outcomes

The extended [36 month] secondary outcomes for this follow-up are also presented in Table 1, by arm, alongside those that were reported at the 12 months follow-up. The predicted means and estimated differences are also given.

Number of cigarettes smoked per day was lower in the BSC group at 36 months than it was at 12 months, and slightly higher in the UC group than at 12 months. There was no evidence of a difference between the arms at 36 months for number of cigarettes smoked per day – IRR 0.92 (95% CI 0.78 to 1.07, p = 0.27).

The FTND scores were similar between the arms at 36 months (mean: 5.8 BSC, 5.7 UC), and were similar to those reported at the one-year follow-up (mean: 5.6 BSC, 5.3 UC). When comparing the total FNTD score at 36 months there was no evidence of a difference between the arms (AMD -0.00, 95% CI -0.48 to 0.47).

For the MTQ there was a noticeable difference in means at 12 months (13.0 BSC and 12.3 UC), and when compared at 12 months there was a statistically significant result. At 36 months the average scores are consistent with the mean scores at 12 months (mean 13.1 BSC, 12.5 UC). When comparing the arms, there was an indication of a different between the arms, but it was not statistically significant (AMD 0.68, 95% CI -0.10 to 1.46, p = 0.09).

The PHQ-9 was recalculated as the PHQ-8 for baseline, 6months and 12 months, and a model rerun for this score, to allow for a comparison with 36 months. The mean scores were similar at 12 months and 36 months, and there was no evidence of a difference between the arms at 36 months (AMD 0.09, 95% CI-1.14 to 1.31, p = 0.89).

Similarly, the GAD-7 average scores were similar at 12 months and 36 months, when comparing the arms at 36 months adjusted mean difference 0.41 (95% CI -1.59, 0.77). The average scores were slightly lower in both arms at 36 months compared to 12 months for the physical component of the SF-12, but similar for the mental component. When comparing the results at 36 months both components were found to be similar between the arms (AMD -0.11, 95% CI-2.34 to 2.11 and -0.39, -2.01 to 1.22 for physical and mental respectively).

The average BMI was around 31 for both arms at 36 months – a slight increase in both arms from 12 months (30.4 intervention, 29.7 control). This now classifies the average BMI as ‘obese’ for both groups at 36 months. There was no evidence of a difference in BMI between the arms at 36 months, adjusted mean difference -0.03, 95% CI -1.34 to 1.29.

## Discussion

The main finding of the SCIMITAR+ trials is that smoking cessation can be achieved in the short term amongst people with severe mental illness.^18,19^ Compared to usual care, the provision of a bespoke smoking cessation intervention increased engagement and the chances of successful quitting as estimated by a biochemically-verified outcome measure. The chances of successful quitting at six months post-randomisation amongst those who received bespoke smoking cessation were more than twice those who received usual care. At 12 months this difference was attenuated, but by 36 months there was no evidence of a difference.

This finding is in line with research in the wider population which shows that long-term quit rates are difficult to achieve and this remains a challenge in the treatment for nicotine dependence in any population.^36^ We also found that there was no enduring between group differences on measures of mental health, including depression and anxiety. This provides supportive evidence that offering a smoking cessation intervention is not detrimental to mental health, but that sustained quitting is difficult to achieve.

SCIMITAR+ was a pragmatic trial^23^ and our comparator was therefore usual care. As such the treatment that is offered under conditions of usual care will be variable, and will often fall short of the ideal or that recommended in evidence supported guidelines.^37^ Previous research has shown that the uptake of smoking cessation services by people with severe mental illness is lower than that for the wider population,^13^ and this has also been found to be the case under conditions of usual care in the SCIMITAR+ trial.^37^

SCIMITAR+ is the first full-scale UK RCT of a combined behavioural and pharmacological intervention designed for people with severe mental illness. Trials to date have been small scale and with short periods of follow up, and have focussed on pharmacological treatments, with limited consideration of behavioural approaches.^21^ The present trial extends research in this area by helping to demonstrate the limited longer-term impacts and to highlight the need for continued monitoring of smoking status and the provision of further offers of smoking cessation to ensure quitting is maintained.

The absence of a clear effect in the longer term is subject to two important caveats. First, the rate of follow up was quite low, even with concerted attention to follow up and efforts to contact participants. There was high levels of missing data [>50%] and it is difficult to assume what might have happened among the participants who were not captured in extended follow up. We assumed that those lost to follow up had in fact continued smoking and this may not be realistic. The low levels of follow up and make it difficult to be confident in establishing longer term impact. Second, the high levels of loss to follow up have significantly reduced the statistical power to detect an effect even if one were present, and there is a risk that a type 2 error is present.

We adapted and enhanced an evidence-supported smoking cessation strategy that has been developed and forms the mainstay of successful UK stop smoking services.^38^ This structured intervention was delivered by a mental health professional and a ‘cut down to quit’ approach was also offered.^39^ The results of the SCIMITAR+ trial, alongside trials^40^ and systematic review evidence ^12,41^of the safety and effectiveness of pharmacological treatments for nicotine dependence in people with mental illness represents accumulating evidence of the effectiveness of smoking cessation interventions for this disadvantaged group. There is also emerging evidence that smoking cessation can be delivered in inpatient settings,^42,43^though the SCIMITAR+ trial specifically excluded such populations. Further research is needed to examine if the SCIMITAR+ intervention could be adapted to be delivered in inpatient environments where patients often experience abstinence for the first time.

The intervention described in the SCIMITAR+ trial (alongside other such models of care^44,45^) now form the template for mental health services in the UK and form the basis of NICE guidance in this area.^46^ On the basis of these results we suggest that clinical services should ensure longer term follow up in order to maintain the short term quit rates observed in the SCIMITAR+ trial and to anticipate high levels of relapse.

In the face of significant health inequalities for people with severe mental illness, smoking is the most important modifiable risk factor for poor health and reduced life expectancy.^46,47^ In the SCIMITAR+ trials programme we have shown that people with severe mental illness more readily engage with a bespoke intervention which results in increased six month quit rates, but that longer term quitting is attenuated and likely to be more difficult to achieve. Health systems should provide smoking cessation interventions that are responsive to the needs of people who use mental health services. Further research is needed to establish how long-term quitting can be supported and maintained.

## Data Availability

Data sharing: reasonable requests for patient level data should be made to the corresponding author and will be considered by the SCIMITAR+ trial management group.

## Contributions of the authors

SG and EP wrote the original SCIMITAR trial protocol. TB, CH, LH, TH, SK, ML, DO, EP, SP, JR and SG, were co-applicants on the HTA application. CA, DB, DBr, TB, SC, CF, CH, LH, MH, PH, TH, SK, JL, ML, DO, SP, JR and SG refined the protocol. SG was the chief investigator and over saw the study. EP was the trial manger. IC and CF conducted the clinical analysis, and CH oversaw the conduct of the analysis. JL designed and undertook the economic analysis in conjunction with SP. DB supervised the delivery of the intervention and CA, SC, TM, PH and TS were trial coordinators for the study. TM, TS, EB, PB, SB, DBr, TC, AC, CC, DC, ED, KE, HH, WK, LN, EN, HO, JRe, CBRH, KS, AS and CV recruited participants to the study, provided feedback on the recruitment methods and helped refine the study procedures. PP and SR helped refine the study procedures. The views and opinions expressed herein are those of the authors and do not necessarily reflect those of the Department of Health.

## Acknowledgements

We would like to thank: the participants for taking part in the trial, the GPs and secondary and tertiary care staff for recruiting participants to the study and completing trial documentation, the Trial Steering Committee and Data Monitoring and Ethics Committee members for the overseeing the study. We acknowledge the support and advice of Dr Andy McEwan, Director of the National Centre for Smoking Cessation and Training for his advice on the use of evidence supported smoking cessation interventions and their adaptation to people with severe mental illness.

This trial is dedicated to the memory of Professor Helen Lester (1961-2013) who collaborated on the early stages of the SCIMITAR+ trial, and is a celebration of her work and contribution to the care and wellbeing of people with severe mental illness. This was her abiding passion and will be her lasting contribution.

## Funding acknowledgment and disclaimer

This trial was funded by NIHR Health Technology Assessment Programme (project number or ref 11/136/52). SG, SP and ML was funded by the NIHR Collaboration for Leadership in Applied Health Research and Care Yorkshire and Humber (NIHR CLAHRC YH) www.clahrc-yh.nihr.ac.uk. The views expressed are those of the authors and not necessarily those of the NHS, the NIHR or the Department of Health and Social Care.

### Data sharing

reasonable requests for patient level data should be made to the corresponding author and will be considered by the SCIMITAR+ trial management group.

---

**Supplementary Table 1:** Baseline characteristics of participants who completed the three-year follow-up, and those who were initially randomised, by arm.

|  | Randomised |  | Followed-up at Y3 |  |
| --- | --- | --- | --- | --- |
|  | Intervention<br>(n=265) | Control<br>(n=261) | Intervention<br>(n=129) | Control<br>(n=132) |
| **Gender, n (%)** |  |  |  |  |
| *Male* | 159 (60.0%) | 150 (57.5%) | 69 (53.5%) | 66 (50.0%) |
| *Female* | 105 (39.6%) | 111 (42.5%) | 60 (46.5%) | 66 (50.0%) |
| *Transgender* | 1 (0.4%) | 0 (0.0%) | 0 (0.0%) | 0 (0.0%) |
| **Age (years)** | N=265 | N=261 | N=129 | N=132 |
| Median | 47.6 | 46.6 | 48.6 | 48.5 |
| IQR | 35.6 – 55.2 | 36.5 – 53.8 | 35.6 – 56.1 | 40.0 – 54.2 |
| **Body mass index (kg/m<sup>2</sup>)** | N=263 | N=258 | N=129 | N=129 |
| Median | 29.0 | 29.4 | 30.1 | 26.1 |
| IQR | 25.1 – 34.0 | 24.9 – 33.3 | 26.1 – 34.4 | 26.1 – 33.6 |
| **Most recent diagnosis** |  |  |  |  |
| *Bipolar Disorder* | 59 (22.4%) | 56 (21.5%) | 34 (26.4%) | 27 (20.5%) |
| *Schizoaffective Disorder* | 25 (9.5%) | 41 (15.7%) | 15 (11.6%) | 21 (15.9%) |
| *Schizophrenia* | 138 (52.5%) | 125 (47.9%) | 65 (50.4%) | 67 (50.8%) |
| *Other psychotic disorder* | 41 (15.6%) | 39 (14.9%) | 15 (11.6%) | 17 (12.9%) |
| **Cigarettes usually smoked (per day)** | N=265 | N=261 | N=129 | N=132 |
| Mean (SD) | 24.7 (13.5) | 23.2 (12.8) | 25.1 (12.5) | 23.7 (12.4) |
| **Smoking duration (years)** | N=265 | N=260 | N=129 | N=131 |
| Mean (SD) | 30.7 (13.2) | 29.0 (12.5) | 31.9 (13.0) | 30.3 (12.4) |
| **Exhaled CO (ppm)** | N=264 | N=258 | N=129 | N=130 |
| Mean (SD) | 24.9 (15.4) | 24.3 (15.1) | 25.0 (14.4) | 24.1 (13.9) |
| **Alcohol consumption** |  |  |  |  |
| *Yes* | 141 (53.2%) | 140 (53.6%) | 74 (57.4%) | 64 (48.5%) |
| *No* | 122 (46.0%) | 121 (46.4%) | 54 (41.9%) | 68 (51.5%) |
| *Missing* | 2 (0.8%) | 0 (0.0%) | 1 (0.8%) | 0 (0.0%) |
| **Do you feel that smoking has affected the state of your health?** |  |  |  |  |
| *Yes* | 220 (83.0%) | 219 (83.9%) | 107 (83.0%) | 114 (86.4%) |
| *No* | 45 (17.0%) | 42 (16.1%) | 22 (17.1%) | 18 (13.6%) |
| **Advised to quit smoking by doctor** |  |  |  |  |
| *Yes* | 192 (72.5%) | 181 (69.3%) | 95 (73.6%) | 97 (73.5%) |
| *No* | 73 (27.5%) | 80 (30.7%) | 34 (26.4%) | 35 (26.5%) |
| **Recreational drug use** |  |  |  |  |
| *Yes* | 20 (7.5%) | 25 (9.6%) | 4 (3.1%) | 8 (6.1%) |
| *No* | 244 (92.1%) | 234 (89.7%) | 125 (96.9%) | 122 (92.4%) |
| *Missing* | 1 (0.4%) | 2 (0.8%) | 0 (0.0%) | 2 (1.5%) |

